# Liver function tests and fibrosis scores in a rural population in Africa: estimation of the burden of disease and associated risk factors

**DOI:** 10.1101/19000968

**Authors:** Geraldine A O’Hara, Jolynne Mokaya, Jeffrey P Hau, Louise O Downs, Anna L McNaughton, Alex Karabarinde, Gershim Asiki, Janet Seeley, Philippa C Matthews, Robert Newton

**Author notes:** These authors contributed equally (Geraldine A O’Hara, Jolynne Mokaya). These authors contributed equally (Philippa C Matthews, Robert Newton). CORRESPONDING AUTHOR: Philippa Matthews.

## Abstract

**Introduction:** Liver disease is a major cause of morbidity and mortality in sub-Saharan Africa. However, its prevalence, distribution and aetiology have not been well characterised. We examined liver function tests (LFTs) and calculated liver fibrosis scores in a rural population in Uganda.

**Methodology:** A cross-sectional survey of LFTs was undertaken in 2011 in a rural population cohort in South-Western Uganda. We classified abnormal LFTs based on reference ranges set in America and in Africa. We derived fibrosis scores (AST to Platelet Ratio Index, fibrosis-4, GGT to platelet ratio, red cell distribution width to platelet ratio, and S-index) to evaluate the potential prevalence of liver disease. We collected information about alcohol intake, and infection with HIV, HBV and HCV, to determine the contribution made by these factors to liver inflammation or fibrosis.

**Results:** Data were available for 8,099 participants (median age 30 years; 56% female). The prevalence of HBV, HCV and HIV infection were 3%, 0.2% and 8%, respectively. The prevalence of abnormal LFTs was higher based on the American reference range compared to the African reference range (e.g. for AST 13% vs 3%, respectively). The prevalence of AST/ALT ratio >2 was 11%, suggestive of alcoholic hepatitis. The highest prevalence of fibrosis was suggested by the GPR score, with 24% of the population falling above the threshold for fibrosis. By multivariate analysis, elevated LFTs and fibrosis scores were most consistently associated with older age, male sex, being under-weight, infection with HIV or HBV, and alcohol consumption. Based on population attributable risk, the highest proportion of elevated fibrosis scores was associated with alcohol use (e.g. 64% of elevated S-index scores).

**Conclusion:** Further work is required to determine normal reference ranges for LFTs in this setting, to evaluate the specificity and sensitivity of fibrosis scores, and to determine aetiology of liver disease.

**KEY FINDINGS:** *What is already known?:*

- Liver disease is not well characterised in many parts of sSA despite the high prevalence of chronic viral infections (HIV, HBV and HCV), and potential exposure to hepatotoxins including alcohol, aflatoxins and traditional herbal medicine.
- Non-invasive blood tests for markers of fibrosis are relatively simple and offer a safe route to assess for liver fibrosis, however, their diagnostic accuracy is not well established in sSA.
- Appropriate reference ranges for LFTs are crucial for optimising the sensitivity and specificity of the detection of underlying liver disease.

*What are the new findings?:*

- There is a disparity in the prevalence of abnormal LFTs in our study cohort when comparing two references ranges (American vs. local reference ranges).
- Based on GPR score, there is a high prevalence of liver fibrosis (almost 1 in 4 of this population) and elevated GPR score is associated with older age, male sex, being under-weight, infection with HIV or HBV, and alcohol consumption.
- Alcohol consumption accounted for 64% of abnormal S-index scores, 32% of elevated FIB-4 scores, and 19% of GPR abnormalities.

*What do the new findings imply?:*

- Appropriate reference ranges for LFTs are necessary to contribute to an understanding of the burden and aetiology of liver disease.
- Alcohol, HIV and HBV are risk factors for deranged LFTs and liver fibrosis, with alcohol making the most significant and striking contribution.
- Further investigation is needed to determine other factors that contribute to liver disease in this setting.

## INTRODUCTION

Liver disease causes an estimated 200,000 deaths each year in sub-Saharan Africa (sSA) as a result of liver cirrhosis and hepatocellular carcinoma (1). More than 80% of Africa’s burden of liver disease has been attributed to endemic blood borne virus (BBV) infections, such as HIV, hepatitis B (HBV) and hepatitis C (HCV), alcohol, hepatotoxic medications (including traditional and herbal medicines), non-alcoholic fatty liver disease (NAFLD) and exposure to aflatoxins (1–3). However, the prevalence, distribution and aetiology of liver disease in many parts of Africa have not been well characterised, and the neglect of cirrhosis has recently been highlighted (2). In order to improve screening for liver disease, and to implement appropriate investigations and intervention, we have undertaken a survey of liver function tests (LFTs) together with demographic data for a large rural cohort in South-Western Uganda (4).

LFTs are usually the first approach to evaluation of liver disease (reference ranges and causes of derangement are summarised in Suppl Table 1). In addition, liver synthetic function can be assessed by measuring prothrombin time; platelet production may be decreased in chronic liver disease due to hypersplenism, decreased thrombopoietin levels and bone marrow suppression (5). Abnormal LFTs are often non-specific and can arise transiently in association with many acute illnesses or usage of medications. However, persistent derangement can indicate chronic liver disease, with associated morbidity and mortality (6). The pattern of derangement can sometimes help to establish aetiology – for example AST/ALT ratio >2 is characteristically associated with alcoholic hepatitis (7,8).

Determination of the origin of liver disease and ascertainment of treatment requirements necessitates accurate characteristation of the degree of liver disease. Liver biopsy allows objective grading of fibrosis and can provide information about the likely aetiology of liver disease based on specific changes to cellular architecture. However, biopsy is costly, requires experts to undertake the procedure and analyse samples, and exposes patients to potentially life-threatening risks. Imaging can also be employed to assess fibrosis. Typically, this comprises ultrasound-based techniques, including fibroscan to derive elastography scores. In most low and middle-income settings, evaluation of liver disease currently depends on use of non-invasive (blood) markers, combined with ultrasound and/or fibroscan when available.

Non-invasive fibrosis blood tests are relatively simple and offer a safe route to assess for liver fibrosis, appealing in resource limited settings. Scores of liver fibrosis, such as AST to Platelet Ratio Index (APRI), fibrosis-4 (FIB-4), GGT to platelet ratio (GPR), red cell distribution width to platelet ratio (RPR) and S-index have been derived using liver enzymes (ALT, AST, GGT) in combination with platelet count. However diagnostic accuracy is not well established in sSA and can be influenced by the population being assessed and the nature of underlying liver disease (9–14). GPR has recently been reported as an independent predictor of significant fibrosis in treatment naïve Gambian patients with chronic hepatitis B (CHB) infection (12). However, further studies are needed to determine the specificity and sensitivity of different scores in different settings.

Appropriate reference ranges for LFTs are crucial for optimising the detection of underlying liver disease (15). Application of reference ranges defined in one setting to different populations may lead to either under- or over-estimation of abnormalities (15–17). As well as being dependent on the population being assessed, the distribution of LFTs in any given setting can also be influenced by the type of instrument, reagents used, and the strength of quality assurance (17). Efforts have been made to establish ‘population-specific’ reference ranges (16,18); one example is through the application of cross-sectional data from seven South-Eastern African countries (16). However, such local reference ranges for Africa have been derived from cross-sectional data collected in adults without addressing the potential prevalence of underlying liver disease. Thus, while American reference ranges potentially over-estimate of the burden of liver disease in an African setting, it is also possible that locally derived reference ranges under-estimate the burden (as they are based on thresholds that have been derived from populations in which liver disease is highly prevalent).

We here set out to assess LFTs and fibrosis scores derived from a large, well defined population cohort in rural South-Western Uganda (19). We applied reference ranges set in both America and in Africa (16,20), in order to assess the possible burden of liver disease, highlighting the discrepancies that arise as a result of the difference between thresholds. We derived fibrosis scores to further evaluate the potential prevalence of liver disease in this setting and to estimate the contributions of alcohol and BBVs to the burden of disease.

## METHODS

### Study design and study population

We conducted a cross-sectional study in a rural population in Kyamulibwa, in the Kalungu district of South-Western Uganda as part of the survey of the General Population Cohort (GPC). The GPC is a community-based cohort established in 1989 with funding from the UK Medical Research Council (MRC) in collaboration with the Uganda Virus Research Institute (UVRI) (21). Regular census and medical surveys have been conducted in this population cohort. In 2011, data collection included screening for viral hepatitis and LFTs among 8,145 adults (≥16 years), which we used for this analysis.

### Data collection

Demographic and health history data were collected using questionnaires and standardised procedures described elsewhere (21,22). Blood samples were drawn at home and transported for testing at the Medical Research Council central laboratories in Entebbe. LFTs (serum AST, ALT, ALP, GGT and BR) were measured using a Cobas Integra 400 plus machine, with Roche reagents. Screening for HIV testing was done using an algorithm recommended by the Uganda Ministry of Health, based on initial screening with a rapid test. If the test result was negative, the participant was considered to be HIV negative. If the test result was positive, the sample was re-tested with the rapid test HIV-1 or −2 Stat-Pak. If both tests resulted in a positive result, the participant was diagnosed as HIV positive. If the tests gave discordant results, the sample was further evaluated with the rapid test Uni-Gold Recombinant HIV-1/2. For those samples assessed by all three tests, two positive test results were interpreted as positive, and two negative results were considered negative. HBV surface antigen (HBsAg) testing was conducted using Cobas HBsAg II (2011-08 V10), and those who tested positive were invited for further serologic testing. HCV was tested using a combination of immunoassays followed by PCR, as previously described (23). Normal serum levels of liver enzymes were classified according to the American reference range (ARR, MGH Clinical Laboratories) and Local Reference Ranges (LRR, (16); Suppl Table 1). We excluded individuals ≤19 years from ALP analysis, since elevated ALP can be attributable to bone growth in teenagers.

Data from the full blood count was used to calculate fibrosis scores (mean corpuscular volume, MCV and platelet count). This was collected starting part-way through the 2011 data collection period; the data are, therefore, population-based, although based on only a subset of the whole cohort (n=1,877).

### Calculation of fibrosis scores and AST/ALT ratio

Where data were available (n=1,877), we calculated APRI, FIB-4, GPR, RPR and S-Index. The formulae for calculating these scores are presented in Suppl Table 2, along with the sensitivity and specificity of each, based on previous studies. We used previously established thresholds to suggest the presence of liver fibrosis, as follows: APRI >0.7 (24), FIB-4 >3.25 (25), GPR >0.32 (12), RPR >0.825 (26), S-index >0.3 (27). We calculated AST/ALT ratio; a score >2 has been associated with alcoholic hepatitis (8).

### Statistical Analysis

We analysed data using standard statistical software, Stata/IC 13 (Stata Corporation, College Station, USA) and GraphPad Prism v7.0. We summarised participant baseline characteristics using proportions (%) and these were stratified by sex. We reported prevalence and distribution of LFTs, laboratory markers of fibrosis and elastography scores using descriptive statistics. We reported p-values from chi-square tests, comparing the proportions of each potential risk factor between male and female participants.

We used logistic regression in our univariate and multivariate analyses, using the threshold for significance set at 0.05, to estimate the odd ratios (OR), along with its 95% confidence intervals (95% CI), to identify potential factors associated with abnormal LFTs and liver fibrosis scores, using a forward stepwise approach to develop our multivariate models. We added risk factors that were identified in the age and sex adjusted univariate analysis to the multivariate model. The final multivariate models for each LFT and liver fibrosis score were obtained by excluding variables in the final model until all remaining variables were associated with abnormal LFTs and liver fibrosis scores at the p<0.05 threshold. Once the final multivariate model had been established, variables that were eliminated through this forward stepwise approach were added back to the model and were reported if associated at the p<0.05 threshold, to reduce the effects of residual confounding. Due to the low number of individuals with active HCV infection at the time of the study, we did not include this sub-group in univariate or multivariate analysis. These HCV RNA positive individuals have been described in more detail elsewhere (28). We present results of multivariate analysis in the form of Forrest plots generated using Microsoft Excel. A tabular form of the multivariate analysis containing the adjusted odds ratios (Adj. OR), and 95% CIs are included in the supplementary section of the manuscript.

### Ethics

Ethics approval was provided by the Science and Ethics Committee of the Uganda Virus Research Institute (GC/127/12/11/06), the Ugandan National Council for Science and Technology (HS870), and the East of England-Cambridge South (formerly Cambridgeshire 4) NHS Research Ethics Committee UK (11/H0305/5). All participants provided written informed consent.

## RESULTS

### Characteristics of study population

We analysed complete data for 8,099 participants (Suppl Table 3). Compared to females, there were more males who were HBV positive, (prevalence 3% vs 2%, respectively; p<0.001) and had consumed alcohol in the past 30 days, (40% vs 33%, respectively; p<0.001). More females were HIV positive (9% vs 6%, respectively; p<0.001). Males were more likely to be underweight (31% vs 16%), and females to be overweight (18% vs 5%); p<0.001 in both cases.

### Proportion of population defined as having abnormal LFTs varies according to the reference range that is applied

The proportion of the population falling above the upper limit of normal (ULN) for each parameter is shown in Table 1, with ALT, AST and GGT distributions in Fig 1A-C (full data for all LFTs are shown in Suppl Fig 1). These results highlight the different burden of disease that can be estimated according to the reference range that is applied, with a higher proportion of the population falling above the ULN when the ARR was applied compared to the LRR (Fig 1A, B). Most striking, for AST, 13% of the population had a value that was deemed to be elevated based on ARR, compared to only 3% based on the LRR (Fig 1B). Using the ARR, ALT and BR were significantly more likely to be above the ULN in males than in females, and ALP was more likely to be higher in females (p<0.001 in each case, Table 1). These sex differences were not apparent when the LRR was applied. OR for deranged LFTs and fibrosis scores according to age and sex is shown in Suppl. fig 2.

**Table 1:**
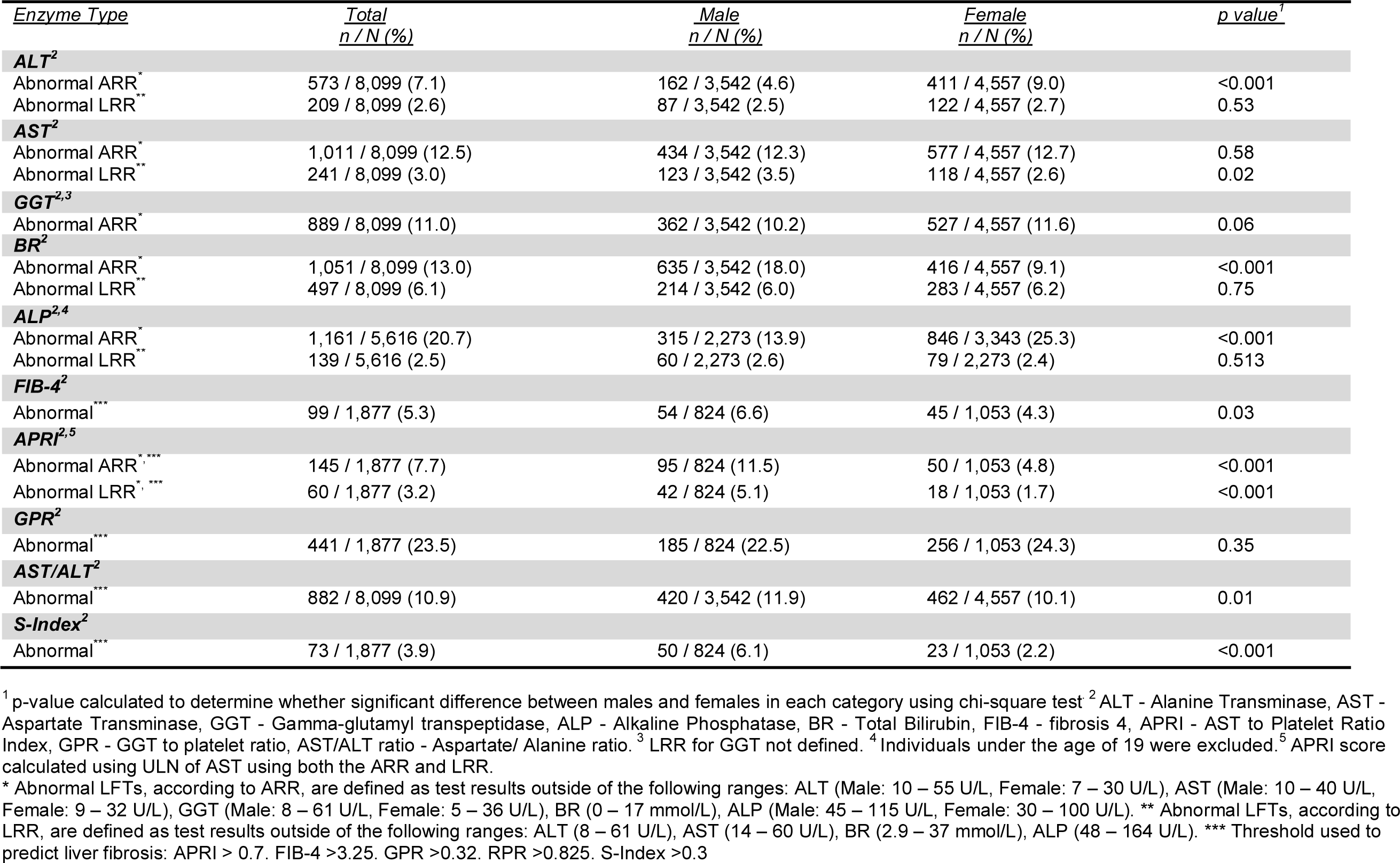
Study participants from the Uganda General Population Cohort with abnormal LFT results and fibrosis scores based on upper limit of normal according to American reference range (ARR) and local reference ranges (LRR).

**Fig 1:**
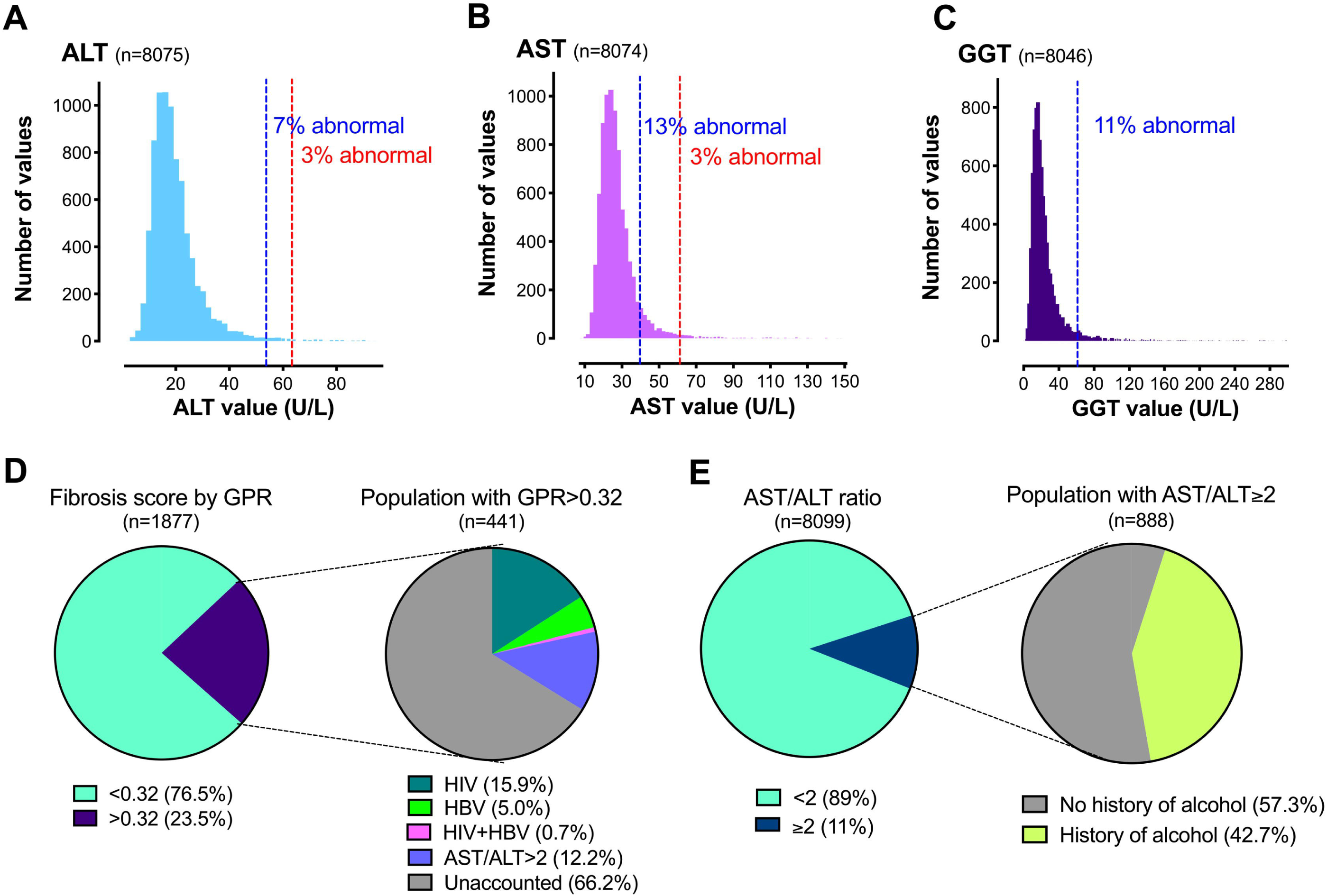
Liver function tests and hepatic fibrosis scores among adults in the Uganda General Population Cohort. Distribution of (A) ALT, (B) AST and (C) GGT. Dashed vertical lines indicate upper limit of normal (ULN) based on American reference range, ARR (blue) and local reference range, LRR (red), as shown in Suppl Table 2. Note no LRR for GGT. (D) Proportion of the population with an elevated GPR score, and among those with elevated GPR the proportion with a defined risk factor for fibrosis. (E) Proportion of the population with an elevated AST/ALT ratio, and among those with an elevated ratio the proportion with a self-reported history of alcohol intake.

### The highest prevalence of liver fibrosis is predicted using the GPR score

We calculated APRI, FIB-4, GPR, RPR and S-index scores (Table 1). The estimated prevalence of fibrosis was highest when based on GPR score (23.5%; Fig 1D), compared to FIB-4 (5.3%), APRI (3.2%), S-index (3.9%) and RPR (0.1%). We excluded RPR scores from further statistical analysis because so few individuals were classified as having an elevated score (we therefore did not have statistical power to detect any factors associated with abnormal score). Because the APRI is derived using the ULN of AST, the proportion of the population classified as having a score consistent with liver fibrosis changes according to whether the ARR or LRR is used (Table 1). Based on previous validation among African individuals, there is evidence to suggest that GPR is the most accurate score for staging liver fibrosis (12); applying this approach, there is a prevalence of almost 1 in 4 adults with liver fibrosis in this population.

### Evidence for the contribution of alcohol to liver disease

The prevalence of AST/ALT ratio >2, suggestive of alcoholic hepatitis, was 11% (888/8,099) (Fig 1E). There was a significant relationship between self-reported alcohol consumption and elevated AST/ALT ratio (p<0.001; Suppl Fig 3). However, 57% of participants with AST/ALT ratio >2 reported never having consumed alcohol (Fig 1E), possibly reflecting either under-reporting of alcohol use and/or other factors that underpin this pattern of LFTs. Self-reported alcohol consumption was associated with raised LFTs, as follows: ALT (Adj. OR 1.33, 95% CI 1.09,1.63) AST (Adj. OR 1.53, 95% CI 1.30, 1.78) GGT (Adj. OR 2.00 95% CI 1.69, 2.36), and with abnormal fibrosis scores, particularly GPR (Adj. OR 1.96, 95% CI 1.52, 2.54). All ORs, adjusted ORs, their respective 95% confidence intervals and p-values are shown in Table 2, and selected variables in Fig 2.

**Fig 2:**
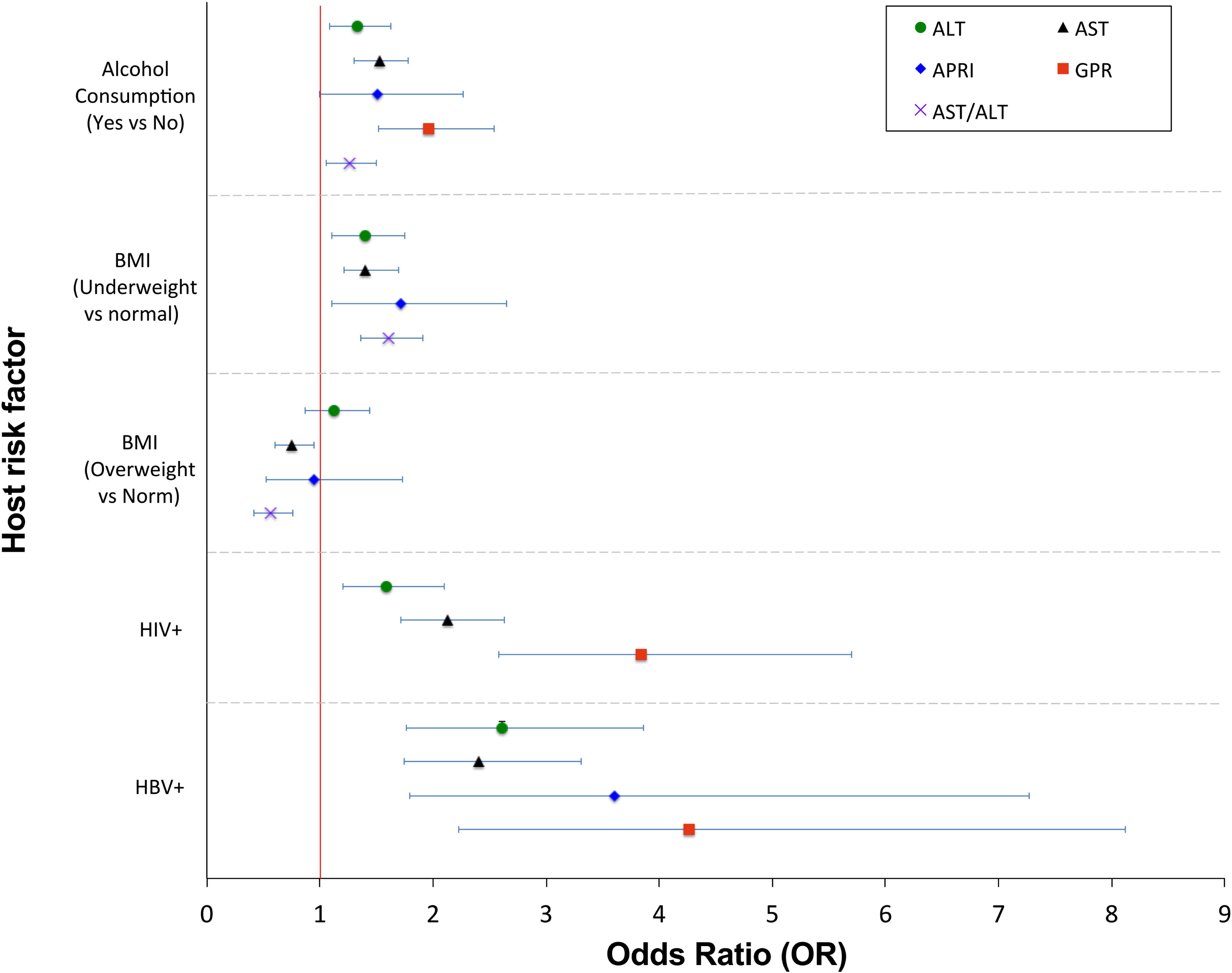
Forrest plots to show odds ratio (OR) for host risk factors and elevated LFTs or fibrosis scores in the Uganda General Population Cohort. Data are presented for the final multivariate model for ALT, AST, APRI, GPR, and AST/ALT and we show variables that were independently associated with the outcome (statistically significant at the P<0.05 level after adjusting for other variables).

**Table 2:**
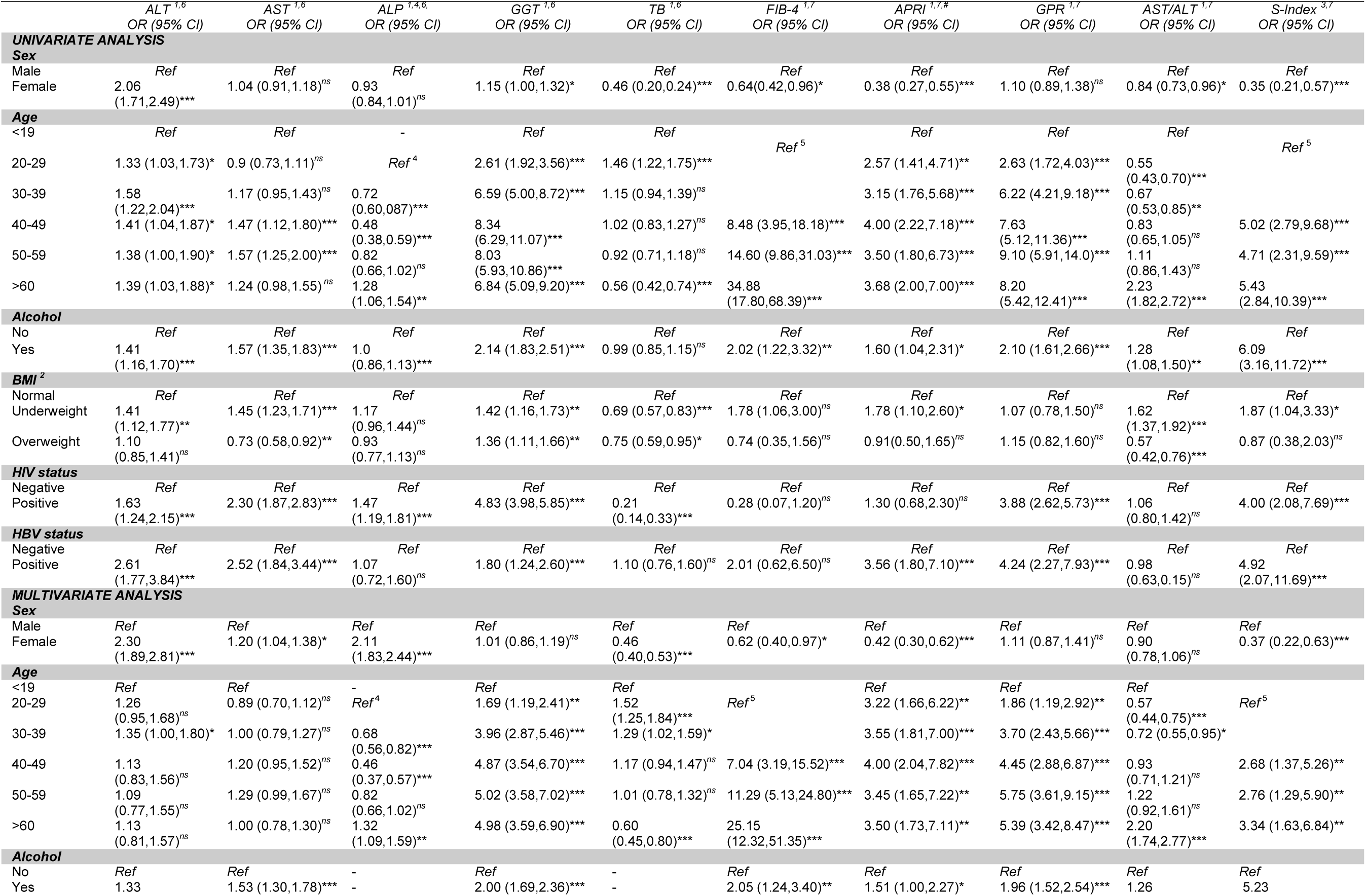
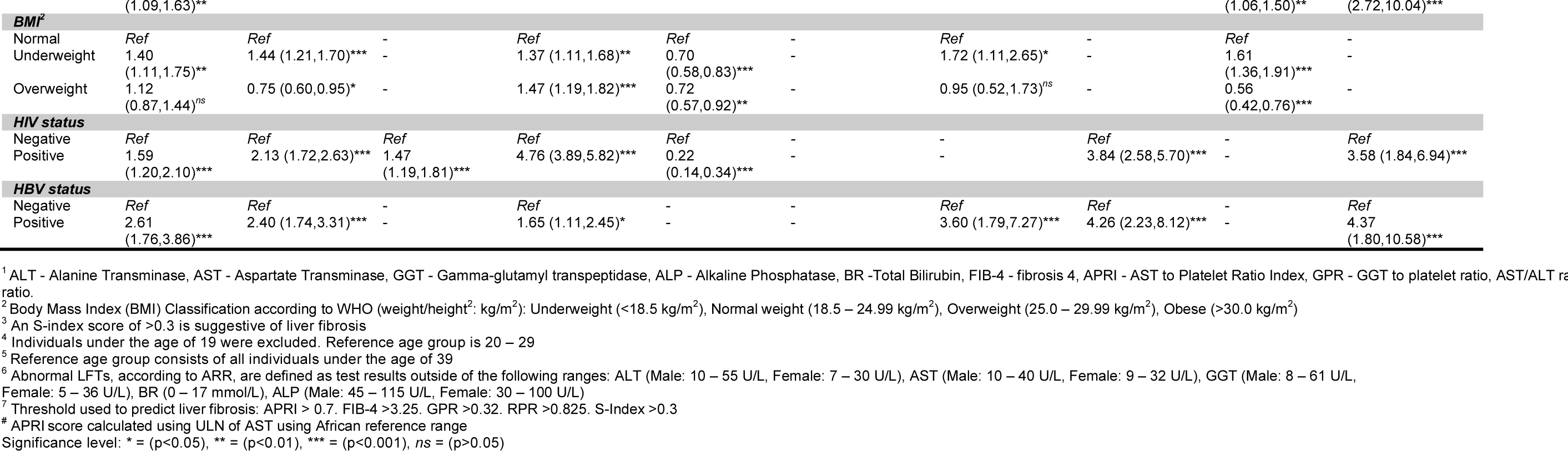
Univariate and multivariate analysis for factors associated with abnormal liver function tests according to American reference rang (ARR) for ALT, AST, ALP, GGT, and TB, and laboratory markers of fibrosis in adults in the Uganda General Population Cohort.

A raised GGT level in combination with AST/ALT ratio >2 can be used to increase the sensitivity of detection of alcoholic hepatitis (8). GGT levels were significantly higher among males with AST/ALT ratio ≥2 (p<0.001), but there was no relationship between GGT and AST/ALT ratio in females (p=0.7); Suppl Fig 4. This potentially indicates that alcohol is of more influence as a cause of an elevated AST/ALT ratio in men than in women. There was no significant association between AST/ALT ratio ≥2 and the presence of an elevated GPR score, predicting fibrosis (p=0.2; data not shown). We calculated population attributable risk (PAR) as a way to assess the relative contribution of different risk factors to the overall burden of liver disease; Table 3. Overall, the most striking contribution arose from reported alcohol consumption, which accounted for 64% of abnormal S-index scores, 32% of elevated FIB-4 scores, and 19% of GPR abnormalities.

**Table 3:**
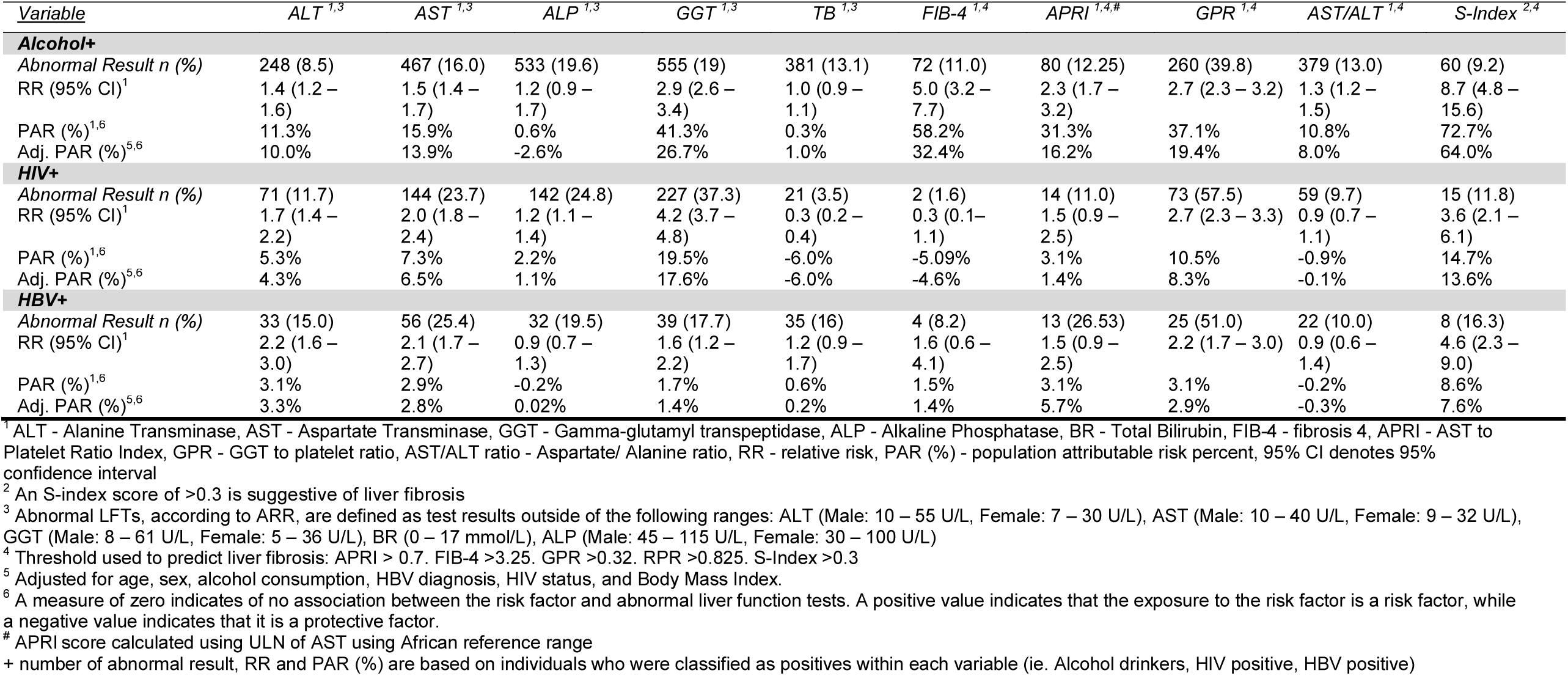
Relative risk, population attributable risk (PAR) percent, and the number of individuals with abnormal liver function tests in the Uganda General Population Cohort. Analysis according to American reference ranges (ARR for ALT, AST, ALP, GGT, and TB)

### Abnormal LFTs and/or elevated fibrosis scores are associated with sex, age, and body mass index

Females were less likely to have high fibrosis scores based on FIB-4 compared to males (Adj. OR: 0.6), APRI (Adj. OR: 0.42), and S-Index (Adj. OR: 0.37) compared to males. FIB-4 score increased markedly with age: adults aged 40 – 49 (Adj. OR: 7.04), 50 – 59 (Adj. OR: 11.29), and adults >60 years (Adj. OR: 25.15) were more likely to have a higher FIB-4 than individuals < 39 years. Elevated BMI was associated only with a rise in GGT (Adj. OR: 1.47). However, being underweight was associated with a more pronounced pattern of liver derangement, including elevations in ALT (Adj. OR: 1.40), AST (Adj. OR: 1.44), GGT (Adj. OR: 1.37), abnormal fibrosis scores (APRI Adj. OR: 1.72,) and with raised AST/ALT ratio (Adj. OR: 1.61). 95% CI in each case are shown in Table 2.

### Relationship between BBV infection and liver disease

HIV infection was associated with abnormal liver function tests, with significant OR for increased ALT, AST, ALP and GGT, as well as with raised GPR and S-index (on univariate and multivariate analysis; Table 2). HBV infection was significantly associated with a rise in hepatic transaminases (OR for raised ALT and AST 2.6 and 2.4 respectively, on multivariate analysis), and with liver fibrosis as measured by APRI and GPR (OR 3.6 and 4.2 respectively, on multivariate analysis). We investigated the prevalence of BBV infection among individuals with raised fibrosis scores. There was an association between the presence of HIV or HBV and raised GPR (p=0.005) and S-Index (p<0.001). Therefore, GPR and S-Index may be the most sensitive markers of inflammation and/or fibrosis in the context of HBV or HIV infection. HIV and HBV were associated with a lesser proportion of liver disease than alcohol based on calculation of PAR (Table 3), but still contributed to elevations in both LFTs and fibrosis scores. The OR for deranged LFTs/fibrosis scores in the context of HIV or HBV infection is shown in Fig 2.

### Liver disease of unknown aetiology

Among individuals with GPR>0.32, 33.8% had either BBV infection or had AST/ALT>2 (suggesting potential alcoholic hepatitis) (Fig 1D; Suppl Fig 5). However, this illustrates that 66% have raised fibrosis scores in the absence of a history of alcohol use, or HIV or HBV infection, suggesting that other factors unaccounted for in this study are likely to be contributing to the overall burden of liver disease. True prevalence of liver disease cannot be ascertained until reference ranges have been more carefully defined, correlating LFTs and fibrosis scores with the confirmed presence of underlying liver disease based on imaging or biopsy.

## DISCUSSION

Liver disease is not well characterised in many parts of sSA despite the high prevalence of HIV and HBV, and potential exposure to hepatotoxins (1,3). In this study, we used cross-sectional data from a large population cohort to estimate the burden of liver disease and to assess the possible impact of BBV infection and alcohol consumption. The prevalence of abnormal LFTs depends on the reference range that is applied. The ARR suggests a higher prevalence of liver disease, therefore including more false-positives. The LRR was established based on individuals recruited from several countries across Africa (Rwanda, Uganda, Kenya, Zambia) (16). While the values were derived from purportedly healthy adults, it is impossible to rule out a high background prevalence of underlying liver disease; in defining higher values for the ULN of all tests, the LRR is more susceptible to false-negatives if used to screen for liver disease.

LFTs are a blunt tool for assessment of liver health, with many potential confounding factors. This current study only accounts for a limited range of aetiological agents, and we did not include other potentially relevant factors such as Schistosomiasis infection, exposure to aflatoxin and use of traditional medications. Furthermore, LFTs were measured at only one point in time, potentially overcalling liver disease as a result of transient abnormalities.

Composite fibrosis scores have been developed with the aim of improving sensitivity of detection of liver disease (29), but these scores also depend on platelet count which can be influenced by diverse factors. For example, in some African populations, thrombocytopenia is common due to infections such as malaria, schistosomiasis, HIV or endemic parasites, as well as being influenced by inflammatory conditions and certain drugs (9,10). We only had platelet counts for a sub-set of our study population, limiting the number for whom we could determine APRI, FIB-4, GPR, S-Index and RPR scores. Data surrounding the use of these scores in sSA is variable, but since in many low-income settings alternative diagnostic equipment is unavailable, non-invasive approaches are vital to estimate liver damage and to stratify clinical management decisions.

APRI and FIB-4 are currently recommended by the World Health Organisation (WHO) for assessment of hepatic fibrosis in patients with chronic HBV or HCV infection (30,31). However, there is evidence showing that APRI is more accurate in assessing liver fibrosis among individuals with chronic HCV compared to HBV infection (11). GPR and S-Index have been validated in small studies in sSA, and have been associated with improved classification of liver fibrosis in chronic HBV infection when compared to APRI and FIB-4 (12–14). It is apparent that either larger studies, or indeed a meta-analysis, are required to further assess the accuracy of these tests in different populations. GPR and S-index may be worthwhile options to include in routine clinical practice to assess for liver fibrosis in African populations, given the high burden of HBV in this continent (32,33). RPR has been used to detect fibrosis among individuals with chronic HBV in China (26), however this score was excluded from our analysis due to a very small number of individuals falling above the suggested threshold for fibrosis.

The prevalence of AST/ALT ratio >2 in this population is 11%, suggesting potential alcoholic hepatitis (34), concordant with a previous study in Uganda in which 10% of the population was estimated to have alcoholic hepatitis (35), and with data from Uganda’s non-communicable diseases risk factor survey which estimated that almost 10% of Ugandan adults have alcohol use disorders (36). Data from emergency attendances at Mulago Hospital in Kampala recorded 47% who reported alcohol use, while 21% and 10% met the study definitions of alcoholic misuse and alcoholic liver disease, respectively (35). Our data are based on self-reported alcohol consumption so may underestimate the true extent of alcohol use. We were unable to quantify alcohol intake or the nature of the alcohol consumed: this is challenging as alcohol is often home-brewed or home-distilled from locally grown grains or fruits, and the alcohol content may vary widely; e.g. the alcohol content of locally produced maize-based brews and liquor in Kenya ranged from 2%-7% and 18%-53%, respectively (36). The global challenge of morbidity and mortality associated with alcohol use is highlighted by recent studies from the Global Burden of Disease consortium, in which alcohol ranks as the seventh highest cause of DALYs and deaths and worldwide (2), and together with HBV infection is a leading aetiological agent of liver cancer (37).

Abnormal LFTs are common in HIV infection for diverse reasons including direct cytopathic effects of HIV on the hepatocytes, co-infection with other BBVs, opportunistic infection, malignancy, ART or other drugs, or secondary to other factors such as alcoholism (38–41). Although a proportion of our study population with fibrosis were infected with BBV (21.6%) and/or had a history of alcohol consumption (12.2%), there was a residual proportion with scores suggestive of fibrosis and AST/ALT ratio >2 who cannot be accounted for through either alcohol or BBV infection. This implies that other factors contribute towards liver dysfunction in this population; a recently published article reported approximately 30% of liver cirrhosis in Africa are not attributed to HBV, HCV, or alcohol misuse and could be as a result of other understudied factors such as NAFLD and use of traditional medicine (35). Aflatoxin exposure is associated with liver cirrhosis and is among the major causes of hepatocellular carcinoma globally, with most cases reported from sSA. Within a previous study of the GPC, >90% of individuals had evidence of exposure (42–44).

In our population women were significantly more likely to be overweight women than men. This may be associated with a higher incidence of NAFLD in women. However, typically only mild rises in ALT are seen, and 80% of those with NAFLD have normal LFTs (45–47) so may not be identified within our current dataset. Diagnosis of NAFLD therefore depends on ultrasound scan (USS); previous studies have consistently shown 70-80% of obese patients have NAFLD on imaging (46,48,49). These imaging modalities were not available in our population, so we are unable to comment specifically on the possible prevalence of NAFLD. Interestingly, in this setting low body weight was more associated with deranged LFTs and with biochemical evidence of liver fibrosis, suggesting a range of pathology that may contribute to liver disease, including organ-specific effects of under-nutrition or stunting (37), as well as the effect of general systemic illness. Further studies are required to investigate the specific relationship between BMI and liver fibrosis in African populations.

In African populations, HCV infection has frequently been often over-reported due to a reliance on HCV-antibody (HCV-Ab) testing, which detects not only current infection but also previous exposure, and is known to be susceptible to false positive results (28). In this cohort, 298/8145 (3.7%) individuals tested HCV-Ab positive, but among these only 13 were HCV RNA positive (overall prevalence 13/8145 = 0.2%).

Appropriate reference ranges for LFTs are necessary to contribute to an understanding of the burden and aetiology of liver disease. Further work is required to determine appropriate thresholds for the ULN of different parameters in different settings in sSA, and to determine which fibrosis score is most specific, through application of a more widespread approach to elastography and/or other imaging. At present, we have identified alcohol, HIV and HBV as risk factors for deranged LFTs and liver fibrosis, with a striking contribution made by alcohol, but further investigation is needed to determine other risk factors that contribute to liver disease in this setting.

## Data Availability

All supporting data are accessible on-line via figshare (URL will be converted to a permanent DOI on acceptance of the paper).

https://figshare.com/s/0b08de8a740991a7aa22

## ABBREVIATIONS

ALT: alanine transferase
APRI: AST to Platelet Ratio Index
ARR: American reference range
AST: aspartate transaminase
BR: bilirubin
BBV: blood borne virus (HIV, HBV, HCV)
FIB-4: fibrosis-4
GGT: gamma glutamyl-transferase
GPC: general population cohort, Uganda
GPR: GGT to platelet ratio
HBV: hepatitis B virus
HCV: hepatitis C virus
HIV: human immunodeficiency virus
LFTs: liver function tests
LRR: local reference range
NAFLD: non-alcoholic fatty liver disease
PAR: population attributable risk
RPR: red cell distribution width to platelet ratio
sSA: sub Saharan Africa
ULN: upper limit of normal
USS: ultrasound scan
WHO: World Health Organisation

## DECLARATIONS

### CONSENT TO PUBLISH

All authors approve the publication of this manuscript

### DATA AVAILABILITY STATEMENT FORMAT GUIDELINES

All data generated or analysed during this study are included in this published article (and its Supplementary Information files).

### FINANCIAL SUPPORT

The General Population Cohort is jointly funded by the UK Medical Research Council (MRC) and the UK Department for International Development (DFID) under the MRC/DFID Concordat agreement. The work on liver function also received additional funding from the MRC, (grant numbers G0801566 and G0901213-92157). JM is funded by a Leverhulme Mandela Rhodes Scholarship. PCM is funded by the Wellcome Trust, grant number 110110. LOD is funded by NIHR.

### AUTHORS’ CONTRIBUTIONS

- Conceived the study : GAO, PCM, RN
- Data collection : AL, GA, RN
- Analysed the data : JM, JPH, PCM
- Wrote the manuscript : GAO, JM, JPH, LOD, PCM, RN
- Revised the manuscript : All authors

All authors have read and approved the manuscript

## ACKNOWLEDGEMENTS

Nil

## SUPPLEMENTARY DATA

All supporting data are accessible on-line via the following link: https://figshare.com/s/0b08de8a740991a7aa22 (this will be converted to a permanent DOI on acceptance of the paper).

**Metadata table:** raw data for 8145 adults in the Uganda General Population cohort (available as .xls and .csv files)

**Supporting data file** (pdf file) contains the following tables and figures:

**Suppl Table 1: Origin, reference ranges and clinical significance of liver functions tests (LFTs)**

**Suppl Table 2: Scores to estimate liver fibrosis, calculated from liver function tests**

**Suppl Table 3: Description of characteristics of study participants with liver function test (LFT) results from the Ugandan General Population Cohort (N=8**,**099)**

**Suppl Fig 1: Distribution of liver function tests in Uganda General Population Cohort**. Dashed vertical lines indicate upper limit of normal (ULN) based on American reference range, ARR (orange line is the ULN for female; blue line is the ULN for males) and local reference range, LRR (black), as shown in Suppl Table 1. Note no LRR for GGT. ULN for bilirubin using ARR is the same for both male and female, indicated by red dashed line. Data are shown for study participants aged ≥16 years, apart from ALP which is shown for participants aged ≥20 to exclude teenagers who may have elevated ALP as a normal physiological consequence of bone growth.

**Suppl Fig 2:** Odds ratio for deranged ALT, AST, APRI, GPR and AST/ALT among participants grouped by sex and age.

**Suppl Fig 3**: **Proportion of Uganda General Population cohort reporting alcohol consumption among individuals with and without AST/ALT ratio >2**

**Suppl Fig 4: Proportion of Uganda General Population Cohort with elevated GGT, according to AST/ALT ratio**. (A) males, with upper limit of normal GGT=61 (B) females, with upper limit of normal GGT=36. P-values by Fisher’s Exact Test.

**Suppl Fig 5: Proportion of Uganda General Population Cohort with blood borne virus (BBV) infection, according to GPR score**. P-value by Fisher’s Exact Test, showing significant enrichment of BBV infection among individuals with elevated GPR score >0.32.

## Notes

CONFLICTS OF INTEREST: We have no conflicts of interest to declare

### Competing Interest Statement

The authors have declared no competing interest.

### Author Declarations

All relevant ethical guidelines have been followed and any necessary IRB and/or ethics committee approvals have been obtained.

Any clinical trials involved have been registered with an ICMJE-approved registry such as ClinicalTrials.gov and the trial ID is included in the manuscript.

